# HOW HEALTHCARE EXPENDITURES AFFECT COVID-19 FATALITY RATE ACROSS EUROPEAN COUNTRIES?

**DOI:** 10.1101/2023.06.23.23291808

**Authors:** Mario Coccia, Igor Benati

## Abstract

The goal of this study is to examine the relationship between healthcare expenditures and health capacity, and variability in COVID-19 case fatality rate between European countries. In particular, the purpose of the present study is to see whether statistical evidence supports the hypothesis that the reduction of COVID-19 fatality, between European countries, can be explained by leveraging health expenditures and if so to form some quantitative analyses and estimates of the relation between health expenditures and COVID-19 fatality rate between countries. The research is based on a sample of European countries and data from various sources, including Eurostat, World Bank, and OECD databases. Results suggest that countries with higher COVID-19 fatality rate in 2020 (when pandemic starts) in comparison to countries with lower COVID-19 fatality had (higher) +50.5% of fatality in 2020, +52.9% in 2022, lower health expenditure as % of GDP −5.5%, health expenditure per capita −34.5%, R&D expenditures in health −30.3%, lower reduction of COVID-19 fatality from 2022-2022 by −57.2 % vs 59.3% of the other group. Results also show a negative association between COVID-19 Fatality in 2022 and Health expenditure as a share of GDP 2020 (*r*=−0.42, *p*-value 0.05); COVID-19 Fatality in 2022 and Vaccinations in December 2021 (*r*=−0.75, *p*-value 0.01). Difference of COVID-19 Fatality 22-20 has also negative correlation coefficients given by *r*=−0.48 (*p*-value 0.05) with Health expenditure as a share of GDP of 2020 and by *r*=−0.52 (*p*-value 0.01) with vaccinations in December 2021. Partial correlation, controlling population over 65yo in 2020, confirms previous results. The contribution here expands the knowledge in these research topics by endeavoring to clarify *how* higher health expenditures improve the preparedness and resilience in crisis management of countries to face unforeseen epidemic or pandemic similar to COVID-19 in society.

## 1. Introduction

The COVID-19 pandemic has affected the whole world since the first months of 2020, driven by manifold environmental, social and economic factors, causing considerable human losses and significant economic and social consequences (Abel and Gietel-Basten, 2020; Bontempi and Coccia, 2021; Bontempi et al., 2021; Chowdhury et al., 2022; Coccia 2020, 2020a,b; Coccia, 2021, 2021a-h; Coccia, 2022f, 2023b; Coccia and Bontempi, 2023; Núñez-Delgado et al., 2021; Tisdell, 2020; Verma and Prakash, 2020). The negative effects of the novel coronavirus were initially countered by states through a variety of non-pharmaceutical measures of control (e.g., lockdown, social distance, facemask wearing, etc.; cf., Akan and Coccia, 2022; Coccia, 2019e,f; Coccia, 2021d,e; Coccia, 2022) and basic health interventions and policies (e.g., care protocols; cf., Amarlou and Coccia, 2023; Ardito et al., 2021; Benati and Coccia, 2022). During the last three years, the effectiveness of the different approaches has been variously discussed and some critical aspects have been clarified (Zhang et al, 2021; Soltesz et al., 2020; Brauner et al, 2020; Bo et al., 2021). Countries that have previously faced the pandemic and experimented with intervention protocols have been able to transfer their know-how to others. In the long run, the intervention models have been standardized and nowadays many aspects of COVID-19 pandemic management appear clearer and more shared (Coccia, 2023, 2023a).

Even though there has been a progressive homogenization of the medical intervention protocols and health policies to combat COVID-19 (Coccia, 2022b), one of the most controversial aspects is to explain why case fatality rates (CFR) of COVID-19 in some countries is higher than other ones (CFR is the proportion of people who die of COVID-19 among all individuals diagnosed with this new infectious disease over a certain period of time). The literature highlights that there are several factors that can contribute to differences in COVID-19 fatality rates across different regions and/or nations (Shakor et al., 2021; Sorci et al., 2020; Khan et al, 2020). These factors are determined by characteristics of population like size and age composition, and health status (Dowd et al., 2020; Sanyaolu et al., 2020; Cao et al, 2020), but also by healthcare expenditure and capacity of countries (Khan et al, 2020; Upadhyay and Shukla, 2021). Cao et al. (2020) found that the size of a country’s population is associated with an increased case fatality rate (CFR) of COVID-19. Age composition is also a relevant factor for negative impact of COVID-19 in society (Coccia, 2020a). Older individuals and those with a lot of comorbidities are at a higher risk of developing severe COVID-19 and dying from this infectious disease. Levin (2020) found that the infection fatality rate (IFR) for COVID-19 is very low for children and younger adults but it increases progressively with age associated with a weaken immune system. Wolff et al. (2020) found that age and comorbidities such as obesity, diabetes, and hypertension are risk factors for severe and fatal disease courses. Sorci et al. (2020) found that the death rate due to smoking in people over 70 years is associated with temporal changes in case fatality rate. In general, countries with older populations may therefore have higher fatality rates associated with pandemic diseases (cf., Galvão et al., 2021).

While demographic factors and characteristics of people are widely investigated, studies that focus on how the healthcare expenditures and other structural factors of socioeconomic systems can affect the impact of COVID-19 in society are hardly know. Some papers suggest that healthcare capacity is associated with the fatality rate for COVID-19. Khan et al. (2020) show that healthcare capacity is associated with a lower case fatality rate for COVID-19. Sherpa (2020) reveals that austerity policies (e.g., health expenditure cuts) significantly increase the mortality rates of COVID-19 in all OCED countries. Other studies suggest that health expenditure is positively associated with case fatalities (Khan, 2020). In this context, Coccia and Benati (2022a) analyze the relationship between public governance and COVID-19 vaccinations during early 2021 to assess the preparedness of countries to timely policy responses to cope with pandemic crises. Results suggest that the improvement of preparedness of countries through good governance can foster a rapid rollout of vaccinations to cope with pandemic threats and the negative effects of their socio-economic impact. To put it differently, countries with better healthcare systems and a greater access of people to medical, such as medical ventilators and personal protective equipment (Coccia, 2023), may be better equipped to manage new airborne diseases, such as COVID-19, in order to reduce fatality rate.

In this research stream, the goal of this study is to examine the relationship between healthcare expenditures and health capacity, and COVID-19 case fatality rate variability in European countries. Healthcare expenditures are intended as the total amount of money spent on healthcare goods and services by individuals, organizations, or governments. These expenditures cover a wide range of healthcare-related expenses, including costs associated with medical services (such as doctor’s visits, hospital stays, and diagnostic tests), prescription drugs, medical devices, public health programs, and health insurance. Healthcare capacity refers to the ability of the healthcare system to provide medical care to individuals who require it. This includes the availability of healthcare facilities, such as hospitals, clinics, and medical equipment, as well as the availability of trained medical personnel, such as doctors, nurses, and other healthcare professionals. The capacity of a healthcare system can be measured in several ways, including the number of hospital beds available, the number of medical professionals in a given area, the amount of medical equipment and supplies on hand, and the overall efficiency of the healthcare system. The choice to refer to the European countries is determined by the need to analyze a homogenous socioeconomic context to minimize variability in the characteristics of the population and the structural indicators. The contribution here expands the knowledge in these research topics by endeavoring to clarify *whether* and *how* health expenditures affect the preparedness and management of countries to face unforeseen pandemic crisis. In particular, this study try to explain the following research questions:

- *What* are countries that have a better preparation to face unforeseen pandemic crisis (e.g., COVID-19) and minimize fatality rates, at beginning when effective drugs and treatments lack?
- *Do* higher health expenditures reduce, associated with other factors, COVID-19 fatality at beginning and during the evolution of pandemic waves?
- *What* are driving forces supporting the resilience of countries to face unforeseen pandemic crisis?

The purpose of the present study is to see whether statistical evidence supports the hypothesis that the reduction of COVID-19 fatality, between European countries, can be explained by leveraging health expenditures and if so to form some quantitative analyses and estimates of the relation between health expenditures and COVID-19 fatality rate between countries. The research is based on a quantitative approach, analyzing data from various sources, including Eurostat, World Bank, and OECD databases. To analyze the effect of healthcare expenditures and capacity on COVID-19 CFRs, we use data from European countries in different time periods, 2020 and 2022. The study utilizes descriptive statistics, *t*-test and regression analysis to investigate whether there is a significant association between healthcare expenditures and capacity and COVID-19 case fatality rate in European countries. Next section describes the study design of this study.

## 2. Study design and methodology

### 2.1 Sample

Our study is based on 27 countries of the European Union (EU) that have a similar socioeconomic structure to have a homogenous sample for robust statistical analyses. In particular, the sample has the following European countries: Austria, Belgium, Bulgaria, Croatia, Cyprus, Czech Republic, Denmark, Estonia, Finland, France, Germany, Greece, Hungary, Ireland, Italy, Latvia, Lithuania, Luxembourg, Malta, Netherlands, Poland, Portugal, Romania, Slovakia, Slovenia, Spain and Sweden.

### 2.2 Variables

This study analyzes 17 variables concerning the characteristics of the population, the resources of the health system and the case fatality rate of the COVID-19 calculated in different years. Table 1 shows the variables under study.

**Table 1.** Variables and sources

| Variable, Acronym, source | Description |
| --- | --- |
| Total population on 1 January 2020, Popolo2020. Eurostat (2023) | The number of persons having their usual residence in a country on 1 January of the respective year. |
| Population 65 years and older on 1 January 2020, Pop65yo2020. Eurostat (2023a) | The number of persons of 65 years or older having their usual residence in a country on 1 January of the respective year. |
| Health expenditure as a share of GDP 2020, HealthExpGDP2020. OECD (2023) | Current expenditure on health (all functions), from all providers and financing schemes (Government and Voluntary) |
| Total health care expenditures per capita in current US\$ 2020, HealthExpPC2020. WHO (2023) | Per capita total expenditure on health (THE) expressed in PPP international dollar. |
| Physicians per 1,000 inhabitants 2018, Physicians1000Inh2018. OECD (2023) | Practicing physicians that provide services for individual patients. It includes: Practicing physicians who have completed studies in medicine at university level (granted by adequate diploma) and who are licensed to practice - Interns and resident physicians (with adequate diploma and providing services under supervision of other medical doctors during their postgraduate internship or residency in a health care facility)<br>- Salaried and self-employed physicians delivering services irrespectively of the place of service provision - Foreign physicians licensed to practice and actively practicing in the country - All physicians providing services for patients, including radiology, pathology, microbiology, hematology, hygiene. |
| Nurses per 1,000 people 2018, Nurses1000Inh2018. OECD (2023) | Practicing nurses that provide services directly to patients. It includes - Professional nurses , Associate professional nurses, Foreign nurses licensed to practice and actively practicing in the country |
| Total hospital beds per 1.000 inhabitants 2019, HBeds1000Inh_2019. OECD (2023) | Total hospital beds are all hospital (HP.1) beds which are regularly maintained and staffed and immediately available for the care of admitted patients. They are the sum of the following categories: a) Beds in publicly owned hospitals; b) Beds in not-for-profit privately owned hospitals; and c) Beds in for-profit privately owned hospitals. |
| Health expenditure in preventive care as a share of GDP 2020, HealthExpPrevCareGDP2020. OECD (2023) | Current expenditure on health (all functions), from all providers and financing schemes (Government and Voluntary) |

Table 1. Variables and sources (continue)
| Variable, Acronym, source | Description |
| --- | --- |
| Hospitals per million inhabitants 2019, HospMillionPeople2019. OECD (2023) | Hospitals (HP .1) comprise licensed establishments primarily engaged in providing medical, diagnostic and treatment services that include physician, nursing, and other health services to inpatients and the specialized accommodation services required by inpatients. It includes: Inclusion - General hospitals (HP 1.1) - Mental health hospitals (HP .1.2). Specialized hospitals (other than mental health hospitals).(HP .1.3) |
| Curative (acute) care beds per 1000 inhabitants 2020. CurativeBeds1000Inh2020. OECD (2023) | Curative care (acute care) beds in hospitals (HP.1) are hospital beds that are available for curative care (HC.1 in the SHA classification). Inclusion - Beds accommodating patients where the principal clinical intent is to do one or more of the following: manage labor (obstetrics), cure illness or provide definitive treatment of injury, perform surgery, relieve symptoms of illness or injury (excluding palliative care), reduce severity of illness or injury, protect against exacerbation and/or complication of illness and/or injury which could threaten life or normal functions, perform diagnostic or therapeutic procedures - Beds for somatic curative (acute) care and psychiatric curative (acute) care (with a breakdown between these two categories) - Beds in all hospitals, including general hospitals (HP.1.1), mental health hospitals (HP.1.2) and other specialized hospitals (HP.1.3) |
| Gross domestic expenditure on R&D in Health in Government sector in US\$ 2020, GrossExpRANDGov2020. OECD (2023) | This table presents data on Gross domestic expenditure on R&D (GERD) by socio-economic objective (SEO), using the NABS 2007 classification i.e.: Exploration and exploitation of the Earth, Environment, Exploration and exploitation of space, Transport, telecommunication and other infrastructures, Energy, Industrial production and technology, Health, Agriculture, Education, Culture, recreation, religion and mass media, Political and social systems, structures and processes, General advancement of knowledge, and Defense. |
| Case fatality rate on 30 December 2020, Fatality 2020. JHU (2023) | The number of deaths in COVID-19 cases divided by the total number of people infected by COVID-19 |
| Case fatality rate on 25 December 2022, Fatality 2022. JHU (2023) | The number of deaths in COVID-19 cases divided by the total number of people infected by COVID-19 |
| Vaccination rate (full cycle) on 26 December 2021, VaccDec 2021. JHU (2023) | The number of people who received two doses of COVID-19 vaccine divided by the total number of population |
| Vaccination rate (full cycle) on 25 December 2022, VaccDec 2022. JHU (2023) | The number of people who received two doses of COVID-19 vaccine divided by the total number of population |

Figure 1 shows the structure of health expenditures in countries and as health expenditures as % of GDP is a wide set including different sub-categories of health expenditures associated with the functioning of overall health system.

**Figure 1.**
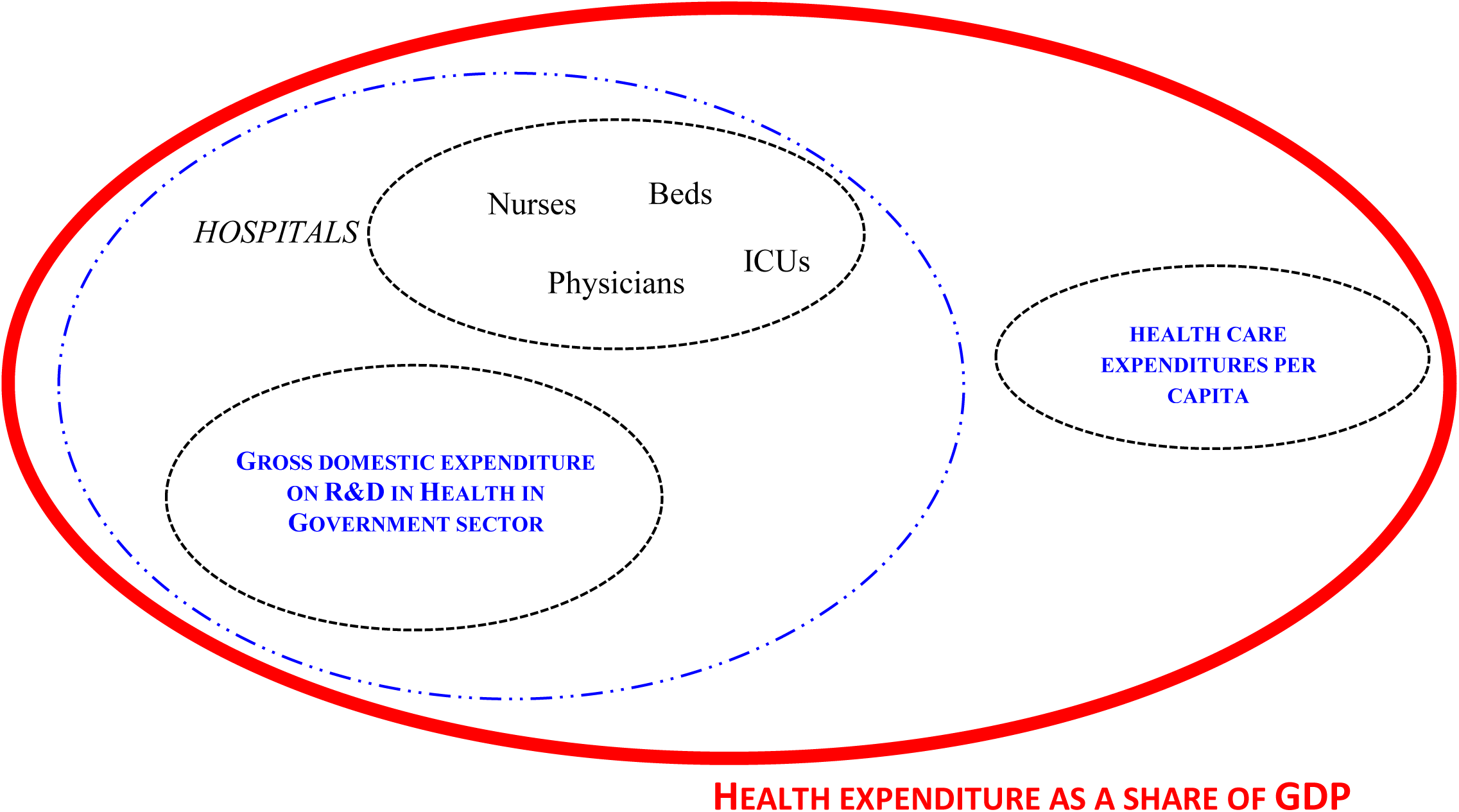
Structure of health expenditure in countries

**Figure 2.**
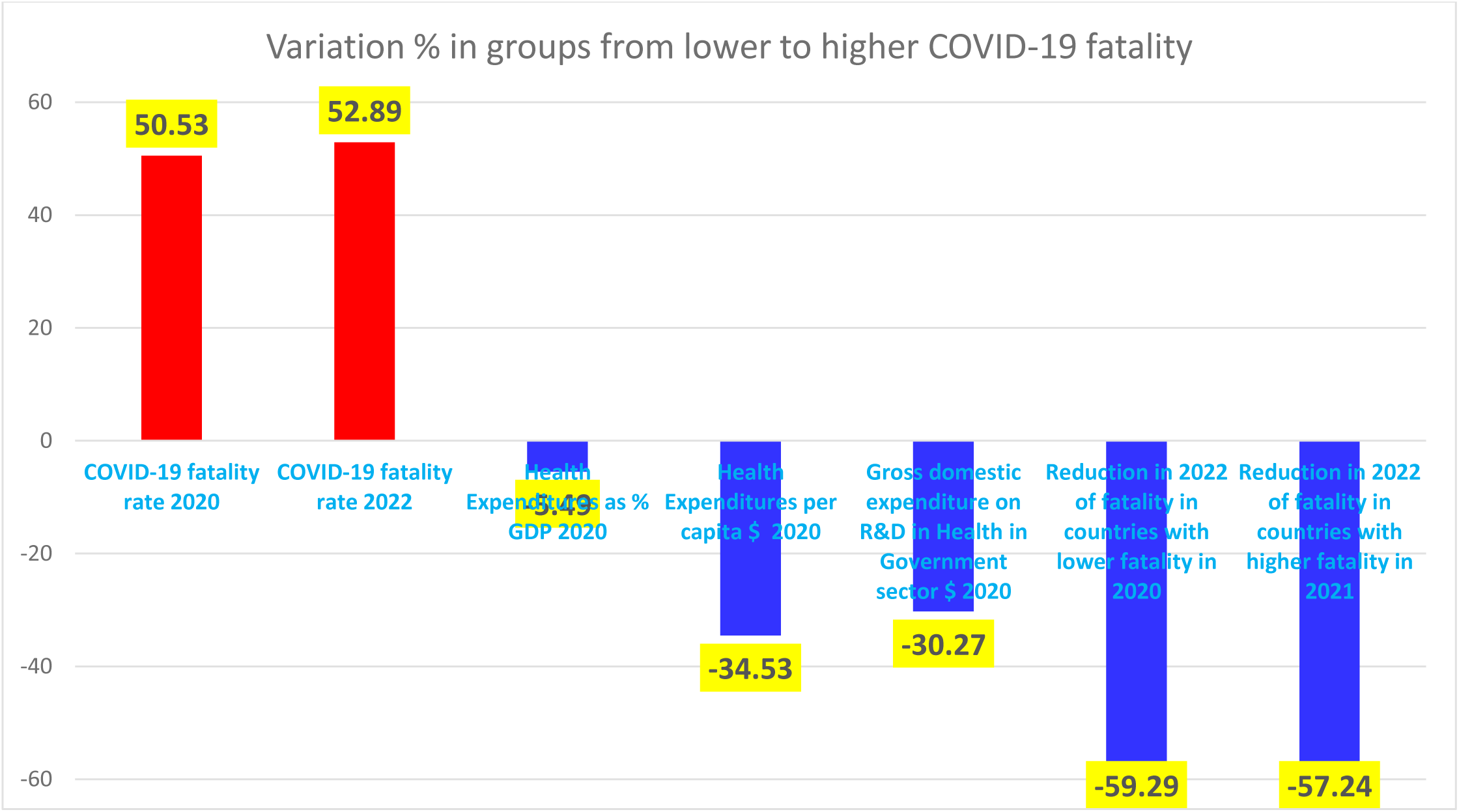
Variation of results between countries with high and low COVID-19 fatality. *Note*: countries with higher COVID-19 fatality rate in 2020 in comparison to countries with lower COVID-19 fatality had (higher) +50.5% of fatality in 2020, +52.9% in 2022, (lower) health expenditure as % of GDP −5.5%, health expenditure per capita −34.5%, R&D expenditures in health −30.3%, lower reduction of COVID-19 fatality from 2022-2022 by −57.2 % vs 59.3% of the other group.

### 2.3 Working hypothesis and Data Analysis Procedure

The purpose of the present study is to see whether statistical evidence supports the working hypothesis that the rate of change of COVID-19 fatality rates in European countries can be explained by the level of health expenditures, and if so to form some quantitative estimate of the relation between COVID-19 fatality rate and health expenditures between European countries.

Firstly, the variables of Table 1 are analyzed with descriptive statistics given by arithmetic mean, standard deviation, skewness and kurtosis to assess the distributions and their normality.

Secondly, the bivariate and partial correlations (controlling population over 65yo in 2020) assess key associations of specific variables under study.

Thirdly, the relationships between some variables are analyzed with a simple regression analysis based on following *log*-*log* model:

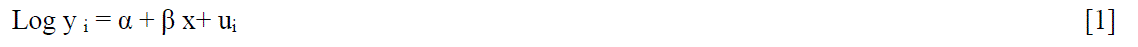

y_i_ = Log COVID-19 Fatality rate December 2020 or Difference in COVID-19 Fatality rate 2022-2020,

*i*=countries

x_i_ = Log Health Expenditure %GDP 2020, *i*=countries

a = constant: the value of response or dependent variable when the associated predictor or independent variable is equal to zero

b = coefficient of regression: it estimates the unknown parameter to describe the relationship between a predictor variable and response variable (Log Health Expenditure %GDP 2020)

Fourthly, average value of COVID-19 fatality rate in 2020 between European countries under study is used to categorize the sample in two groups:

– Group 1A: Countries with a lower COVID-19 fatality rates in 2020 than sample arithmetic mean
– Group 2A: Countries with a higher COVID-19 fatality rates in 2020 than sample arithmetic mean Mutatis mutandis, for COVID-19 fatality rate 2022, the groups are:
– Group 1B: Countries with a lower COVID-19 fatality rates in 2022 than sample arithmetic mean
– Group 2B: Countries with a higher COVID-19 fatality rates in 2022 than sample arithmetic mean

By using average value of the Difference in COVID-19 Fatality rate 2022-2020 between European countries under study, the sample is categorized in the following two groups:

– Group 1C: Countries with a lower value of the Difference of COVID-19 Fatality rate 2022-2020 than sample arithmetic mean; it means a higher reduction of COVID-19 fatality rate in 2022 compared to 2020, suggesting a higher learning process and resilience of countries to cope with COVID-19 pandemic crisis.
– Group 2C: Countries with a higher value of the Difference of COVID-19 Fatality rate 2022-2020 than sample arithmetic mean, it means a lower reduction of COVID-19 fatality rate in 2022 compared to 2020, suggesting a lower learning process and resilience of countries to cope with COVID-19 pandemic crisis.

Finally, data are analyzed with Independent Samples Mann-Whitney U test. The Mann-Whitney U test is used to compare differences between two independent groups when the dependent variable is not normally distributed. The Mann-Whitney U test allows here to draw main conclusions about arithmetic means of groups considering the following null hypotheses:

HP1 A. The distribution of variables is the same across the category of COVID-19 fatality rate in 2022

HP1 B. The distribution of variables is the same across the category of difference in COVID-19 Fatality rate 2022-2020.

Asymptotic significances at the level 0.05 are displayed.

## 3. Empirical analysis

Table 3 shows that a negative association between COVID-19 Fatality December 2022 and Health expenditure as a share of GDP 2020 (*r*=−0.42, *p*-value 0.05); COVID-19 Fatality December 2022 and Vaccinations in December 2021 (*r*=−0.75, *p*-value 0.01); Difference of COVID-19 Fatality 22-20 has also negative correlation coefficients given by *r*=−0.48 (*p*-value 0.05) with Health expenditure as a share of GDP 2020 and by *r*=−0.52 (*p*-value 0.01) with Vaccinations in December 2021.

**Table 2.**
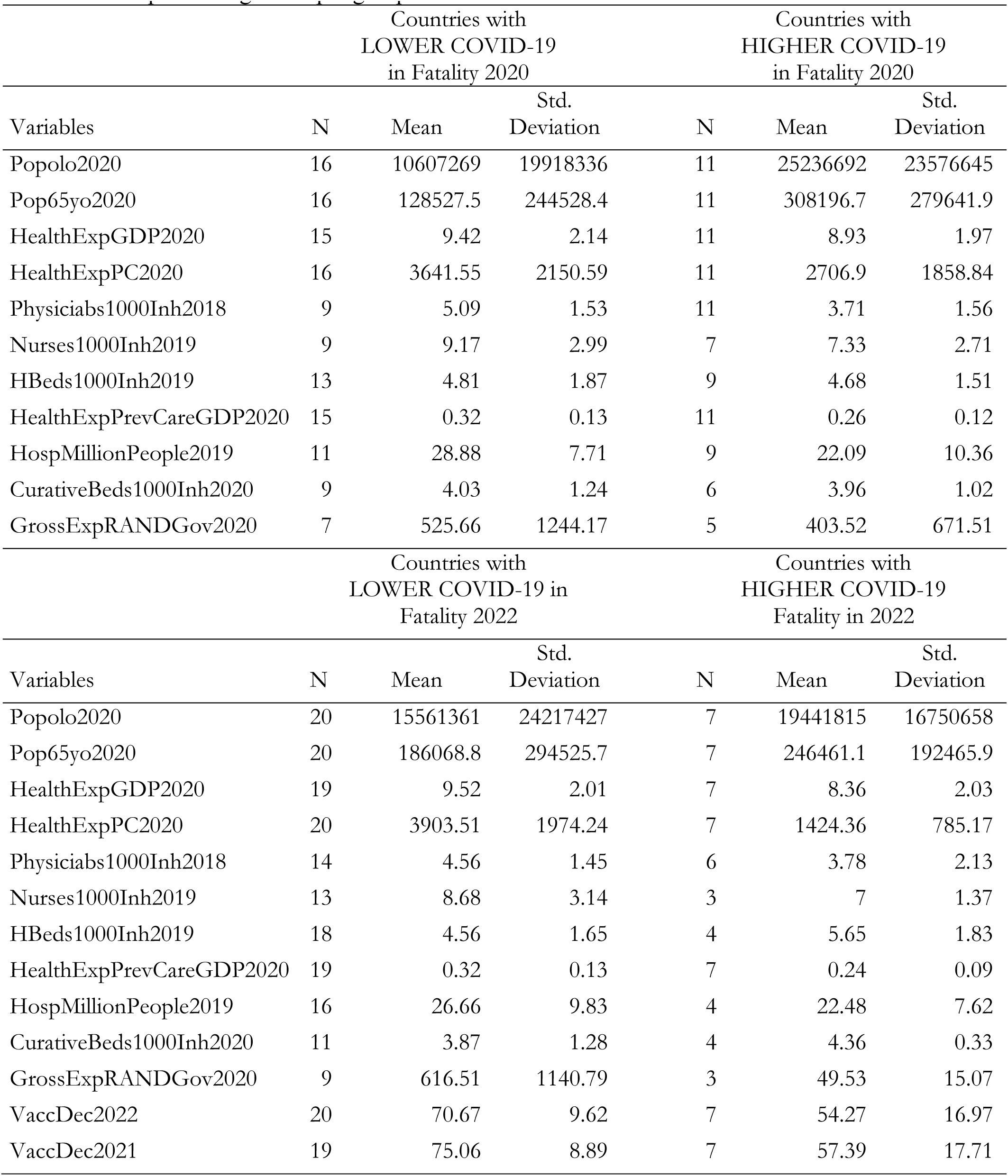
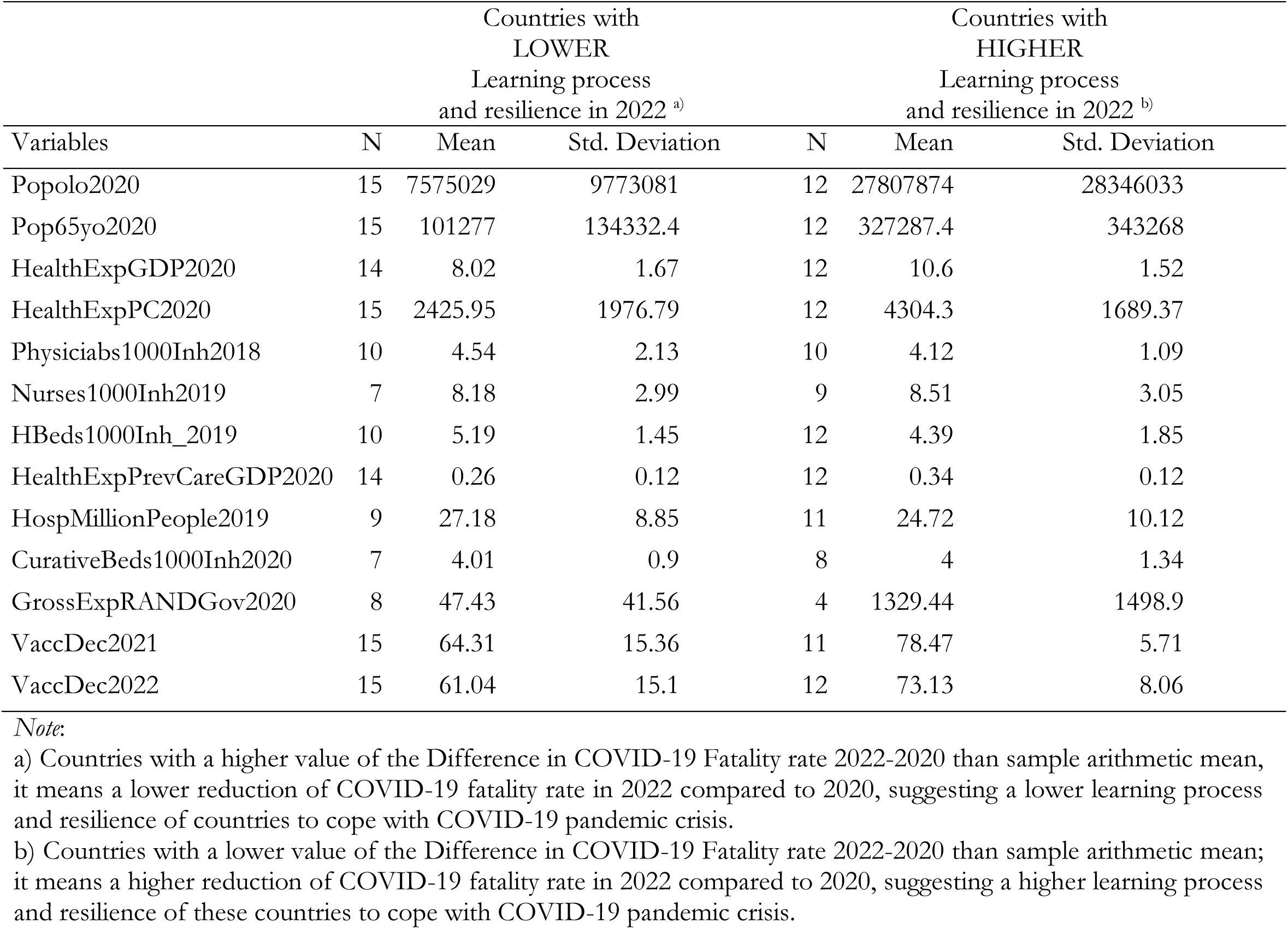
Descriptive categorized per groups

**Table 3.**
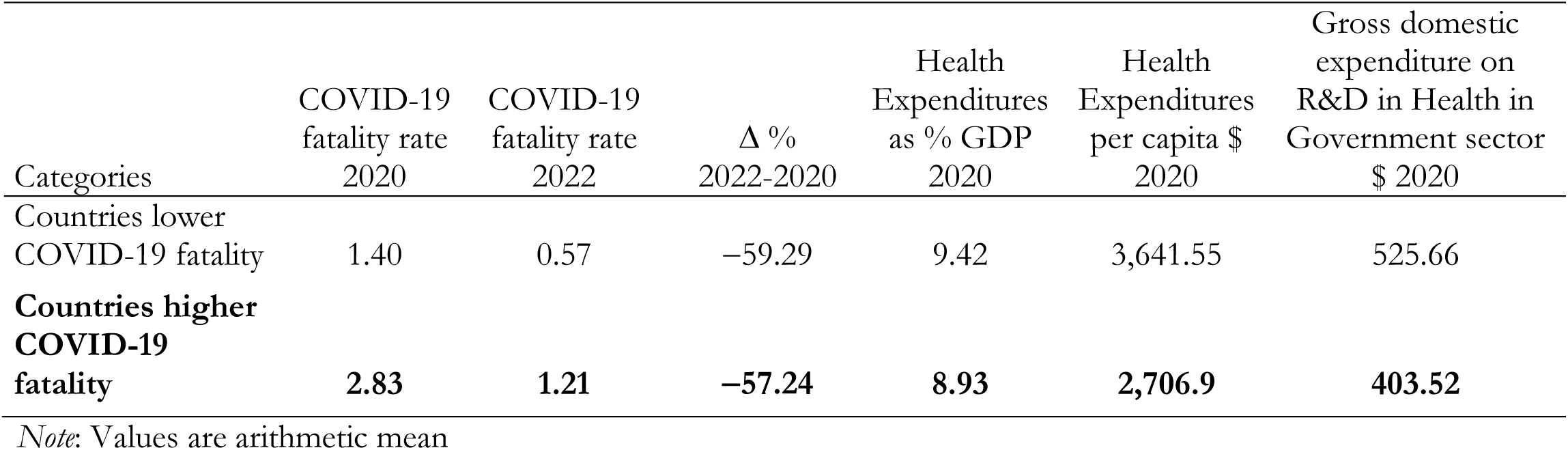
Results between countries with high and low COVID-19 fatality

Table 4 of partial correlation, controlling population over 65yo in 2020, confirms previous results. The role of higher Health expenditure as a share of GDP 2020 and Vaccinations in December 2021 can mitigate COVID-19 Fatality December 2022.

**Table 4.**
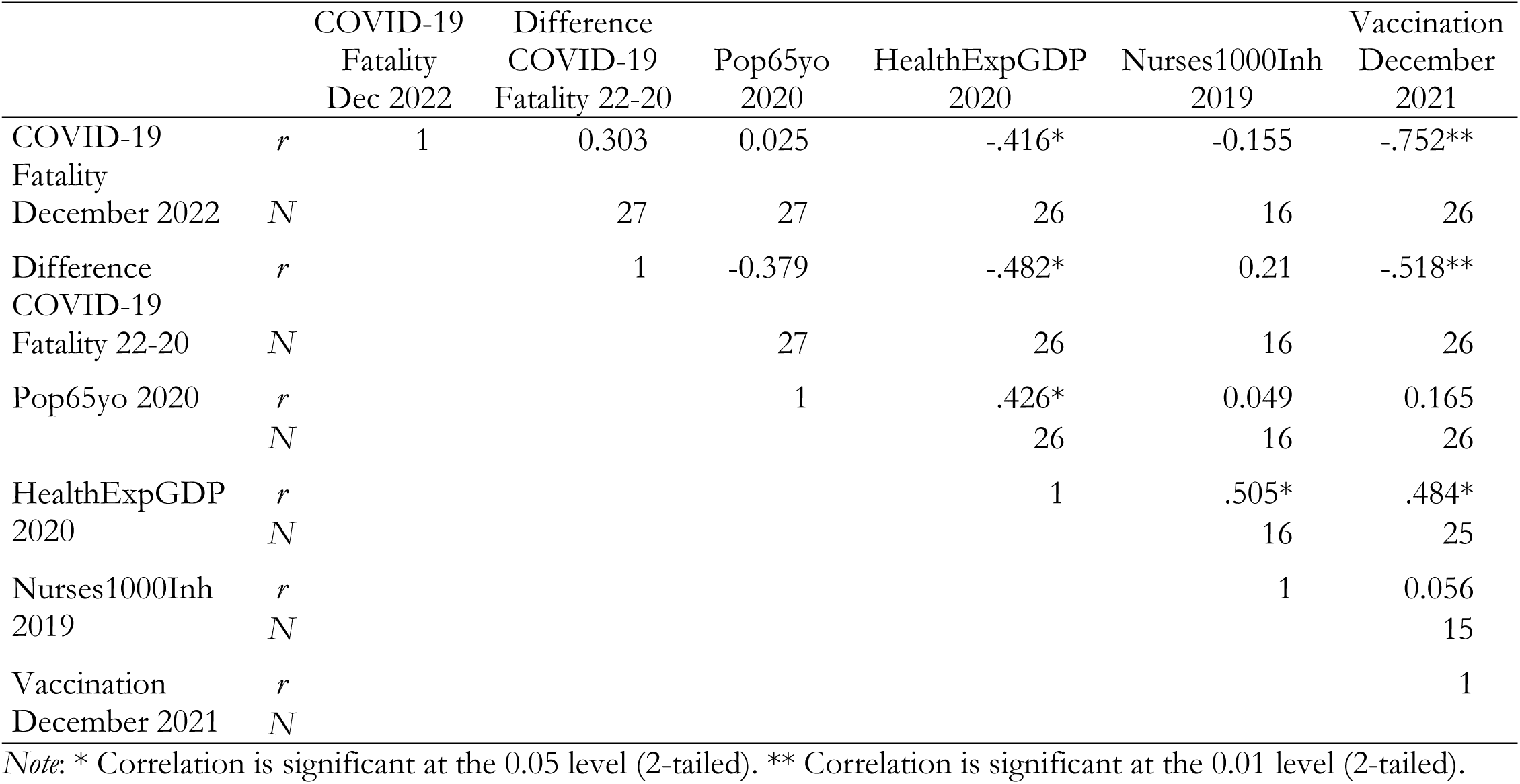
Correlation

Tables 5 shows simple regression analysis: the linear *log-log* model considers the COVID-19 Fatality rate 2020 between European countries as dependent variable (Model 1). Results clearly shows that countries with 1% increase of the Health Expenditure % GDP 2020, they reduce the level of COVID-19 Fatality rate December 2020 by 1.34%. The coefficient R^2^ explains about 17% variance in the data. Although R^2^ is not high in the model, the *F* value is significant (*p*-value<0.05), then independent variable reliably predicts dependent variable (i.e., COVID-19 Fatality rate reduction %). Model 2 in tables 5 considers the Difference COVID-19 Fatality rate 2022-2020 between European countries as dependent variable: it is a proxy of learning processes and resilience to face COVID-19 pandemic crisis (Model 2). Results clearly shows that countries having 1% increase of the Health Expenditure % GDP 2020, these countries reduce the Difference COVID-19 Fatality rate 2022-2020 by 1.84% (*p*-value 0.01). The coefficient R^2^ explains about 23% variance in the data. The *F* value is significant (*p*-value<0.01), then independent variable reliably predicts dependent variable that indicates learning processes between countries to cope with COVID-19 pandemic.

**Table 5.**
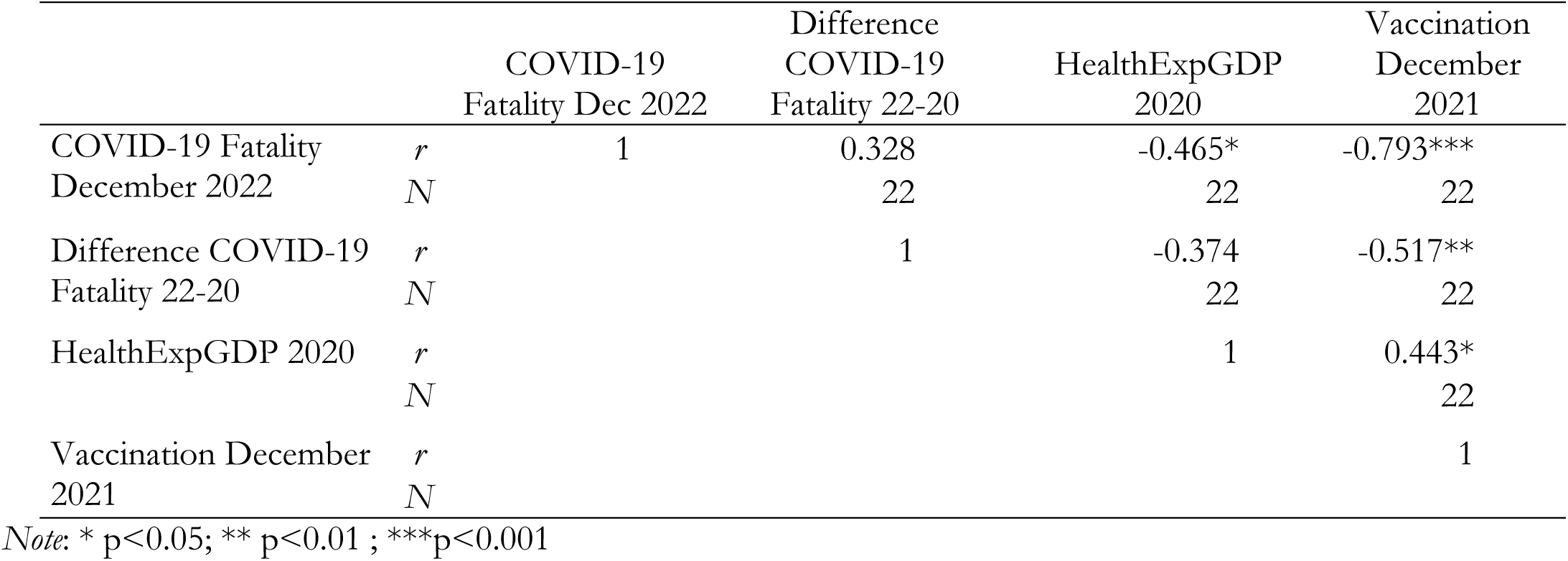
Partial Correlation, controlling population over 65yo in 2020

Finally, table 6 shows Independent Samples Mann-Whitney U test. To compare differences of variables between two independent groups given by countries with lower and higher COVID-19 fatality rate in 2022, and countries having lower and higher difference in COVID-19 fatality rate 2022-2020. Results show that for health care expenditures per capita in current US$ 2020 and vaccinations in 2022 and 2022 the Mann-Whitney U test rejects the null hypothesis that distribution of variables is the same across countries with lower and higher COVID-19 fatality rate in 2022, whereas for the Mann-Whitney U test for health expenditure as a share of GDP 2020, health care expenditures per capita in current US$ 2020, Gross domestic expenditure on R&D in Health in Government sector in US$ 2020 and vaccinations in 2021 and 2022 rejects the null hypothesis that the distribution of these variables is the same across the category of countries with higher and lower difference in COVID-19 Fatality rate 2022-2020. From this data and empirical analyses, it can be concluded that for variables just mentioned COVID-19 fatality rates is significantly lower in groups of European countries with higher health expenditures and level of vaccinations. This group of countries with higher health expenditures has shown a higher preparedness to face COVID-19 crisis for manifold factors including technological, organizational and institutional aspects (Benati I., Coccia M. 2022a; Coccia, 2012, 2012a, 2017, 2017a, 2018; 2019, 2020c-d, 2022a, 2022c-g, Coccia and Benati, 2018; Flaxman et al., 2020; Magazzino et al., 202; Mosleh et al., 2022; Núñez-Delgado et al., 2021, 2023; Yaun, 2020).

**Table 6.**
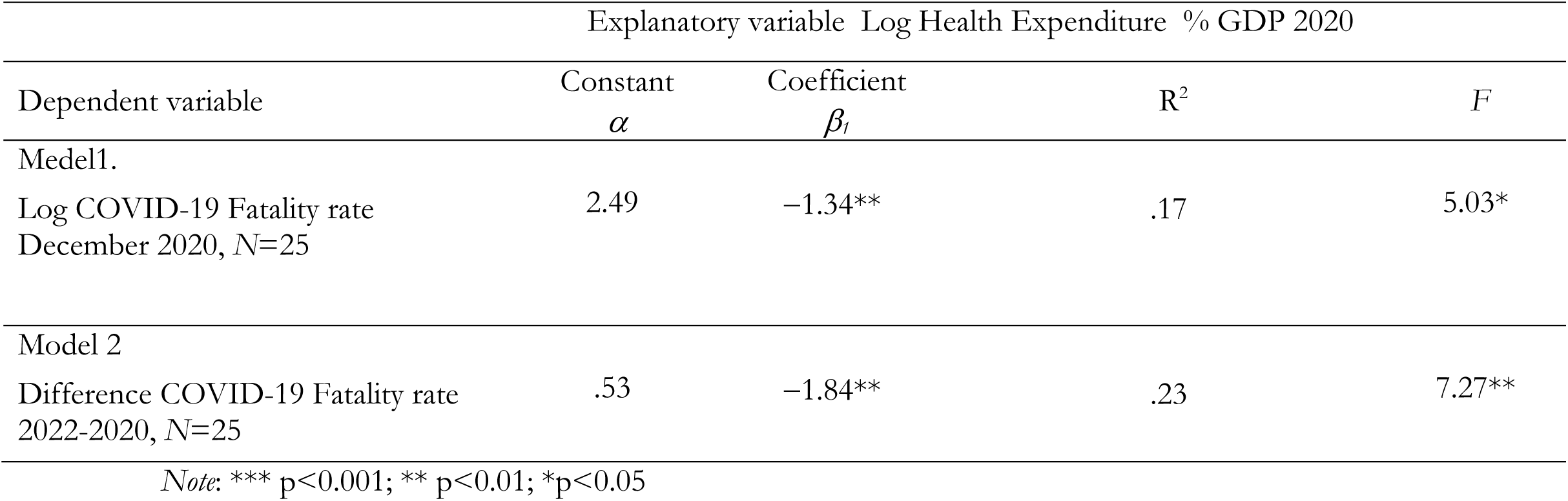
Parametric estimates of the relationship, *log-log* models

**Table 7.**
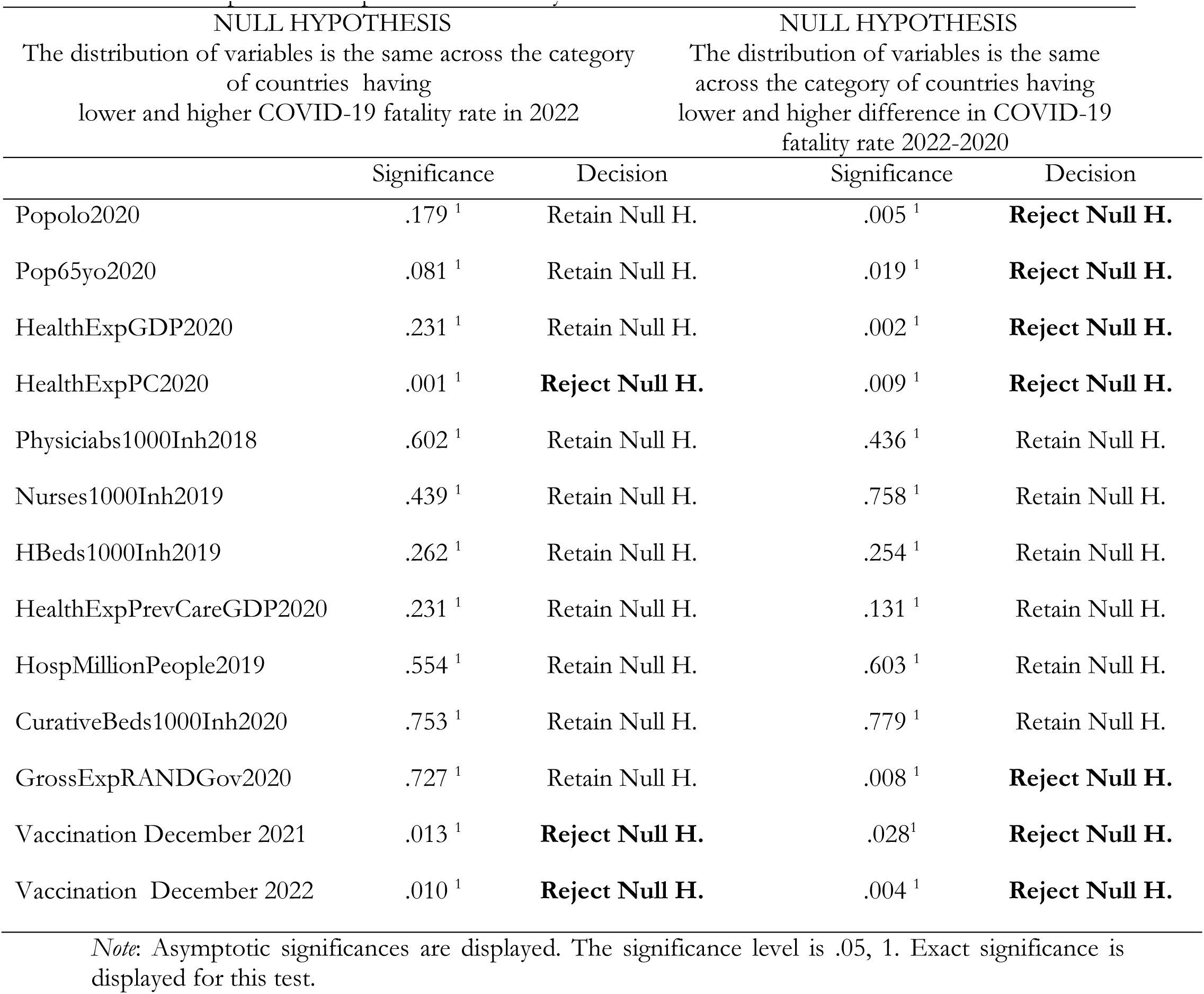
Independent Samples Mann-Whitney U test

## 4. Discussion

The analysis of the data has highlighted the existence of a negative relationship between health expenditure and COVID-19 fatality rate. When the first grows, the second decreases. These results can be due to several factors. Firstly, countries with higher health expenditures generally have better-equipped healthcare systems, which allow for earlier and more accurate diagnosis, more effective treatment, and better health management of crisis (Coccia, 2021, 2021c, 2022c-e). These countries may also have better governance and consequential public health infrastructure, such as disease surveillance and outbreak response systems, which can help to contain the spread of the virus and prevent outbreaks with appropriate health policies (Benati and Coccia, 2022, 2022a). Secondly, higher health expenditures may be associated with better overall health outcomes, including lower rates of chronic diseases and comorbidities that can increase the severity of COVID-19. Countries with stronger healthcare systems may also have higher rates of vaccination and more effective public health contact tracing, which can reduce the spread of novel viral agents and protect vulnerable populations (Benati and Coccia, 2022, 2022a). Finally, higher health expenditures may enable countries to invest in research and development of vaccines and treatments, which can reduce the severity and spread of the COVID-19 and similar infectious diseases (Coccia, 2022c; 2022d, 2022e, 2023a). These countries may also have better access to medical supplies and equipment, such as personal protective equipment (PPE) and ventilators, which can help to mitigate the impact of the pandemic (cf., Coccia, 2023).

Overall, the negative relationship between health expenditure and fatality rate of COVID-19 highlights the importance of investing in healthcare systems and public health infrastructure, supported by a good governance, as a means for improving preparedness in crisis management and protecting populations from the impact of new infectious diseases similar to COCID-19 (Benati and Coccia, 2022, 2022a; Coccia, 2021, 2021c). The analysis of data has also highlighted the existence of a weak relationship between health capacity and fatality rate of COVID-19 (see table 1). This evidence could be attributed to several factors. First, health capacity may not be solely determined by healthcare spending or infrastructure but from overall structure and governance of institutions. Other factors, such as the quality of healthcare delivery, the availability and training of healthcare professionals, and the level of coordination between public health agencies and healthcare providers can also play a role in determining health capacity to face emergencies. Therefore, investing in healthcare infrastructure or increasing healthcare expenditures may not necessarily result in a significant improvement in health outcomes and preparedness if it is not associated with a general good governance of national system. Second, the relationship between health capacity and COVID-19 fatality rate may be influenced by other situational factors such as population density, age distribution, climate, air pollution, etc. (Coccia, 2020, 2020b, 2021b, 2021f). Countries with higher population density and older populations in regions having high air pollution and lower temperature and wind speeds may be more vulnerable to COVID-19 and similar infectious diseases, even if good healthcare systems. Similarly, countries with high rates of chronic disease in population may experience higher COVID-19 fatality rates, regardless of their health capacity. As a last argument, the weak relationship between health capacity and COVID-19 fatality rate, suggested by descriptive statistics, may be due to differences on how countries report COVID-19 deaths. In general, variations in testing capacity, reporting standards, and data collection practices can make it difficult to compare, ceteris paribus, COVID-19 fatality rates across countries. In short, it challenging to identify clear patterns or relationships between health capacity and COVID-19 outcomes. Overall, then, while strong healthcare systems and public health infrastructure are essential factors for protecting populations from infectious diseases like COVID-19, these factors may not be the only determinants of higher or lower fatality rates (Coccia, 2022a). Other factors, such as population characteristics, guidelines in data reporting, climate, air pollution, new technologies, skilled human resources, etc. can also influence the relationship between health capacity and COVID-19 outcomes (Coccia, 2018, 2018a-b, 2019, 2019a-c, 2021l; Coccia and Bellitto, 2018c-d).

## 5. Conclusions

The dynamics and effects of COVID-19 pandemic in society are due to a variety of factors associated with environmental pollution, climate, public governance, institutions, healthcare system, national system of innovation, etc. (Allen Douglas, 2022; Angelopoulos et al., 2020; Ball, 2021; Barro, 2020; Coccia, 2019; Goolsbee and Syverson, 2021; Homburg, 2020; Pagliaro and Coccia, 2021; Wieland, 2020). In the presence of pandemic crises, one of the goals of nations is to mitigate mortality and support the socioeconomic system (Coccia, 2021i). Studies analyze different interventions to cope with the spread of COVID-19 but how level of health expenditures can mitigate negative effects given by infections and deaths of new viral respiratory disease in society is hardly known. The statistical evidence in sections above seems in general to support the working hypothesis stated in Section 2, that the case fatality rates of COVID-19 can be explained by the level of health expenditures between European countries that operate in a homogenous socioeconomic area. In particular, the findings here suggest, between European countries, a negative relationship between health expenditure and COVID-19 fatality rate, such that 1% increase of the Health Expenditure % GDP 2020, it reduces the level of COVID-19 Fatality rate by 1.34%. Moreover, 1% increase of the Health Expenditure % GDP 2020, it reduces the Difference COVID-19 Fatality rate 2022-2020 by 1.84% (p-value 0.01), improving the resilience of countries to face pandemic crises. In short, high levels of investments in healthcare sector, high levels of R&D investments and in new technology of mechanical ventilators, etc. support the preparedness and resilience of countries to face unknown infectious respiratory (cf., Coccia, 2021c, 2023, 2023a). To put it differently, the preparedness of countries for next pandemic crises should be oriented to strengthen health system to cope with future health emergencies, especially when effective drugs to treat patients with acute respiratory illness are missing (Ardito et al., 2021; Kluge et al., 2020; Kapitsinis, 2020). Hence, considering results of the study here, a basic aspect to cope with pandemics is a systematic planning, which should consider continuous investments in health sectors to support IT infrastructure, staffing and training.

Overall, then, results of this analysis here seem to suggest that in the first pandemic wave of COVID-19, countries with a high investment in health sectors and high healthcare expenditure per capita experienced a lower fatality rate of COVID-19. The findings here propose a general strategy of crisis management for future pandemic threats based on high levels of investments in healthcare sector also focused on a widespread implementation of new technologies (such as high-tech medical ventilators) for improving national performance in health emergencies and support the overall preparedness of countries to cope with negative effects of pandemic impact on health of people and socioeconomic system. These conclusions are of course tentative. There is need for much more research in these topics because not all the confounding factors that affect the COVID-19 fatality rates and role of health expenditures and capacity are discussed. Hence, results here have to be reinforced with a follow-up investigation based on new factors of economic systems for additional analyses of the relation between health expenditures and COVID-19 fatality rate to better assess how it can reduce negative effects of next potential respiratory diseases (similar to COVID-19) in society.

## Data Availability

All data produced in the present study are available upon reasonable request to the authors

